# Perceptions of anonymised data use and awareness of the NHS data opt-out amongst patients, carers and healthcare staff

**DOI:** 10.1101/2020.09.12.20193276

**Authors:** C Atkin, B Crosby, K Dunn, G Price, E Marston, C Crawford, M O’Hara, C. Morgan, M. Levermore, S. Gallier, S. Modhwadia, J. Attwood, S Perks, A.K. Denniston, G Gkoutos, R. Dormer, A. Rosser, A. Ignatowicz, H Fanning, E Sapey, On behalf of the PIONEER Data Hub

**Affiliations:** PIONEER Hub in Acute Care, Institute of Inflammation and Ageing, University Hospital Birmingham NHS Foundation Trust, University of Birmingham, Edgbaston, Birmingham, B15 2GW, UK.. ORCiD ID = 0000-0003-0596-8515; PIONEER HDR-UK Data Hub in Acute Care, Institute of Inflammation and Ageing, University Hospital Birmingham NHS Foundation Trust, University of Birmingham, Edgbaston, Birmingham, B15 2GW, UK.; HDR-UK Midlands Physical Site, University Hospital Birmingham NHS Foundation Trust, University of Birmingham, Edgbaston, Birmingham, B15 2GW, UK.; Patient contributor. Patient Involvement and Engagement Lead, PIONEER.; Research Support Services, University of Birmingham, Edgbaston, Birmingham, B15 2TT.; Research and Development, University Hospital Birmingham NHS Foundation Trust, University of Birmingham, Edgbaston, Birmingham, B15 2GW, UK.; University Hospital Birmingham NHS Foundation Trust, University of Birmingham, Edgbaston, Birmingham, B15 2GW, UK.; Public contributor.; a. Chief Executive Officer, Medical Devices Technology International Limited (MDTi), The KaCe Building, Victoria Passage, Wolverhampton, West Midlands, WV14LG, United Kingdom. b. Visiting Professor in Health, Education and Life Sciences, Birmingham City University, Birmingham, West Midlands, UK.; Technical Director, PIONEER HDR-UK Data Hub in Acute Care, Institute of Inflammation and Ageing, University Hospital Birmingham NHS Foundation Trust, University of Birmingham, Edgbaston, Birmingham, B15 2GW, UK.; PIONEER HDR-UK Data Hub in Acute Care, University Hospital Birmingham NHS Foundation Trust, University of Birmingham, Edgbaston, Birmingham, B15 2GW, UK.; Informatics, University Hospital Birmingham NHS Foundation Trust, University of Birmingham, Edgbaston, Birmingham, B15 2GW, UK.; A. Director of INSIGHT - the Health Data Research Hub for Eye Health, University Hospitals Birmingham NHS Foundation Trust, Edgbaston, Birmingham, B15 2GW, UK. B. Centre for Regulatory Science and Innovation, Birmingham Health Partners, Birmingham, B15 2GW, C. NIHR Biomedical Research Centre (Moorfields Eye Hospital NHS Foundation Trust and University College London); Turing Fellow, Alan Turing Institute, HDR-UK Associated Researcher, Institute of Cancer and Genomic Sciences, University of Birmingham, Edgbaston, Birmingham, B15 2GW, UK.; Insignia Medical Systems Limited, Paterson House, Hatch Warren Lane, Basingstoke, Hampshire RG22 4RA, UK; Research Lead, West Midlands Ambulance Service Foundation Trust, Millennium Point, Waterfront Business Park, Waterfront Way, Brierley Hill, West Midlands, DY5 1LX.; Institute of Applied Health Research, University of Birmingham, Edgbaston, Birmingham, B15 2TT, UK.; A. Director of PIONEER, HDR-UK Health Data Research Hub in Acute Care, Birmingham Acute Care Research Group, Institute of Inflammation and Ageing, University of Birmingham, Birmingham, B15,2GW, B. Department of Acute Medicine, University Hospitals Birmingham NHS Foundation Trust, Birmingham B15 2GW. ORCiD ID: 0000-0003-3454-5482, Email; PIONEER Health Data Research Hub, University of Birmingham, Edgbaston, Birmingham, B15 2GW, UK.

**Keywords:** Data sharing, Secondary data use, National Data opt-out, Anonymised healthcare data, Commercial

## Abstract

**Introduction:** Public awareness and support for secondary health data use may vary by health care experience and participant demographics. England provides an example of a centralised “opt out” for secondary use of anonymised health data. We explored the awareness, support for and concerns about anonymised healthcare data secondary use and the NHS data opt-out system amongst patients, carers, healthcare staff and the public within the West Midlands.

**Methods:** A patient and public engagement program was completed, including patient and public workshops, questionnaires regarding anonymised health data use and feedback discussion groups.

**Results:** Central concerns for health data use included unauthorised data re-use, the potential for discrimination and profit generation without patient benefit. Key priorities were projects leading to patient benefit, oversight by the NHS as a trusted organisation, increasing awareness of the NHS data opt-out, and ongoing public/patient involvement.

Questionnaires showed 31.8% were aware of the NHS data opt-out. 93.8% were happy for their data to be used for NHS research, 84.8% for academic research and 68.4% by health companies. However, opinion varied with demographics (age, gender or public, patient, NHS staff and volunteers).

Agreed action points for health data use were education regarding the National Data Opt-Out, public involvement in data requests, NHS oversight, and transparency.

**Conclusion:** Use of anonymised healthcare data for secondary purposes is acceptable to most patients, carers and healthcare workers. However, awareness is limited, and initiatives to publicise potential benefits are needed amongst patients, healthcare staff and the public.

**1) What is already known?:** The secondary use of health data without explicit consent has been widely debated. The potential benefits are clear but public groups have raised concerns, especially when anonymised data is shared with commercial entities.

**2) What does this paper add?:** Perceptions of and support for secondary health data use vary by demographic (age, gender) and experience of health services (Staff member, patient, member of the public). Knowledge of schemes to limit secondary data use (such as the UK National Data Op-Out) are low, even among NHS staff. Patient and public agreed themes to increase the acceptability of health data secondary use include education about ‘Opt-out’ schemes, health service oversight of data use (as the most trusted partner), public and patient involvement in data sharing decisions and public transparency. This framework may increase the acceptability of health data use.

**Strengths:**

1. Mixed methods approach including workshops and questionnaires
2. Includes children aged 13 and over, which is important given they can ‘opt-out’ of health data use at this age using the UK’s National Data Opt-Out.
3. Includes demographics of the diverse participants, rarely collected in most online surveys
4. Includes NHS Staff members, patients and current non-patients, but people with experience of NHS services

**Limitations:**

1. West Midlands based and not national
2. Limited numbers (300+ sample) preventing analysis of sub groups.
3. Participant selection included people with experience of NHS hospital services, and therefore may not be generalisable

## Introduction

The National Health Service (NHS) is a single publicly-funded health service for the United Kingdom (UK) which is free at the point of need to the entire population. The NHS necessarily holds identifiable patients’ medical records within specific NHS organisations^(1)^ but this is confidential^(2)^ and classed as sensitive^(3)^. Electronic health records (EHRs) facilitate data sharing, which is beneficial for the individual^(4)^ especially when care is co-delivered across organisations^(5)^. Also, EHRs facilitate health data sharing for health service planning, research and innovation, termed secondary use^(6)^ but this usually involves anonymised data^(7)^.

Previous research suggests there is support for data sharing^(8)^ but there are public concerns^(2, 9, 10)^, especially where data is not fully anonymised or made available for commercial use^(8, 11)^. This was recently highlighted^(12)^ with UK citizens reporting they were willing to share their data in the following percentages; with academic or medical research institutions (50.3%); a pharmaceutical company (19.8%) or a tech company with an aim to improve health care (12.2%). Little information was provided about the characteristics or healthcare utilisation of the respondents^(12)^ and it is unclear if these preferences might be context dependent - for example if people were frequent users of healthcare services, or if data sharing was overseen by an organisation local to the participant.

The UK’s NHS is a centralised health system in which the secondary use of data is supported but the capability to opt-out exists, although there is some variation across the UK. In England, organisations can apply to use specific data fields from all anonymised patient records, unless patients have ‘opted out’ through the National Data Opt-Out^(13)^. Where patients have not ‘opted-out’, it is presumed that they have no objection to their data being used for secondary purposes, within the limits of national guidance^(2,13,14)^.

Although the National Data Opt-Out is publicised online and physically in healthcare organisations, studies suggest there is variable public awareness of this scheme^(15)^. Suggested frameworks to maximise the secondary use of health data often do not include patients’ and citizens’ views^(16)^ although the importance of these views are widely recognised^(17)^. There is a national consultation to review the Caldicott Principles which guide data use, including consideration of an additional principle that patients’ and service users’ expectations must be considered and informed when confidential information is used^(18)^.

PIONEER is a Health Data Research Hub in acute care, developed to curate routinely-collected health data from unplanned healthcare contacts across community and hospital providers and then facilitate the transparent and ethical use of de-identified data for research and innovation purposes, with a direct aim of improving NHS patient care. PIONEER is based in the West Midlands and to ensure data use reflected the wishes of patients’ whose data are included in the hub, a regional patient and public involvement and engagement (PPIE) programme was initiated, to assess current knowledge and perceptions of health data use.

## Methods

PIONEER is an ethically-approved research database and analytical environment (East Midlands – Derby Research Ethics 20/EM/0158). This PPIE work was conducted following ethical approval from the University of Birmingham Ethical Committee (reference ERN_20-0118).

### Setting and activity

The project included patients and staff from University Hospitals Birmingham NHS Foundation Trust (UHB), one of the largest NHS Trusts in England, with 2750 beds, more than 22,000 staff, a fully EHR (Prescribing Information and Communication System) and a shared primary and secondary care record (Your Care Connected). Members of the public were recruited from public stands and public involvement groups across the West Midlands. For more details of participant recruitment and questionnaire delivery see the online supplement.

The project included six activities, running between February 2019 and July 2020, as shown in Table 1.

**Table 1.** The structure of patient and public activity.

| Activity | Agenda | Objectives | Participants |
| --- | --- | --- | --- |
| 1. Patient workshop | 1. Introduction to health data and electronic health records<br>2. Health data research (benefits and challenges)<br>3. Models of data use (Consent versus unconsented, National Data Opt-Out, broad access for hypothesis generating, limited access and the 5 safes)<br>4. Free discussion – what do attendees think are important considerations in health data use | 1. Understand what is meant by health data and how it is stored for primary use<br>2. Introduce secondary use of health data, de-identification, NHSE opt out and examples of use of health data<br>3. Agree a list of main concerns about health data use<br>4. Agree on a list of important themes to be considered in health data use | Patients who had recently been admitted to UHB |
| 2. Public workshop | 1. Introduction to health data and electronic health records<br>2. Health data research (benefits and challenges)<br>3. Models of data use (Consent versus unconsented, National Data Opt-Out, broad access for hypothesis generating, limited access and the 5 safes)<br>4. Workshop breakout groups and feedback to all<br>a. Key considerations to using<br>patient data<br>b. What processes should guide the use of patient data<br>c. The use of sensitive data | 1. Understand what is meant by health data and how it is stored for primary use<br>2. Consider examples of how patient data has improved patient care but also risks and challenges<br>3. Agree a list of main concerns about health data use<br>4. Agree on a list of important themes to be considered in health data use | Public members from advertisements placed on social media and UHB public forum |
| 3. | Co-creation of questionnaire | Create and test a simple questionnaire to assess knowledge of NHS opt out, and knowledge and perception of health data use. | Patient and public members of PIONEER |
| 4. | Delivery of questionnaire | Delivery of questionnaire to patients, carers, NHS staff and public members including children aged 13 and over | UHB patients, carers and staff. Public members from West Midlands |
| 5. Feedback of questionnaire | Feedback of results of questionnaire to patient working group and public workshop | Assess if wider public consultation reflected the group's thoughts and where differences were present, why these might be present | Patients and public members in two different sessions |

Public members and patients were involved in all stages of the design, delivery, analysis and dissemination of this work.

Participants in the workshops and questionnaire respondents were told that the purpose of the project was to support the development of working practices for PIONEER. Interpreters were used as required.

#### Patient Workshop

An open workshop was held with patients of UHB, who had been admitted to hospital within the past year and were members of a patient support group. This lasted 4 hours with a scribe to take notes. Identified themes were then approved by attendees.

##### Public Workshop

The workshop lasted 4 hours with whole group discussion and smaller break out groups. The workshop and break-out groups were audio-recorded with a scribe to take notes. Themes and action points were recorded and then approved by attendees.

##### Questionnaire co-creation and delivery

A short questionnaire was developed by patient/public members and the PIONEER Health Data Research Hub team. See Table S1 of the online supplement for the questions asked. Verbal consent was given by all participants and where children were approached, their parent or responsible adult provided verbal consent.

##### Feedback sessions

Participants at the patient working group and public workshop were invited to a further meeting where the results of the questionnaire were presented and discussed. In a free discussion, participants considered thse results and formed agreed founding principles for data access processes. The workshops were audio-recorded with a scribe to take notes. The identified principles were then approved by attendees.

#### Data analysis

Stakeholder workshop audio-recordings were transcribed. Notes of group discussions were also reviewed. As data was collected, thematic analysis was undertaken in an iterative process, searching for commonly expressed views, feelings or words. Summaries and initial themes of the workshops and working groups were shared with participants for their feedback. Participants commented on the findings and particularly on any areas that they felt had been misunderstood. They were also encouraged to make further comments and agree themes.

Demographic data from the questionnaire (age, sex, ethnicity) was compared to the 2011 Birmingham census data and Jan 2019 - Jan 2019 UHB patient episode data. Statistical analyses were performed using IBM SPSS statistics. Comparisons between groups were performed using Chi squared and Fisher Exact tests. A p value of <0.05 was considered statistically significant.

#### Results

##### Patient workshop

Twelve Birmingham-based patients were included with a median age of 68 years (range 39 - 78), 58.3% identified as female, the rest as male. They had a median of 3 admissions to hospital in the last year (IQR 2 - 3.5) and all had chronic respiratory diseases. Five members of the group described themselves of Black, Asian or minority ethnic (BAME) background (41.7%) and seven as of White ethnic group (58.3%).

There was unanimous agreement that routinely collected health data could improve healthcare, with recent examples discussed^(19)^. The agreed, key concerns about health data use were;

1. Unauthorised data re-use or sharing.
2. Re-identification of the individual
3. Data used to discriminate against groups or the individual.
4. Data being used to generate commercial profit which did not benefit the NHS or UK population.

A key area of discussion was the question of who controlled access to the data. The participants agreed it was important that a trusted partner oversaw data access and use, with the NHS identified as the most trusted partner. There was agreement that data minimisation was the most appropriate model for access to unconsented health data; with a defined project and data fields, and in accordance with the ‘5 safes’^(20)^.

83.3% people present had not heard of the National Data Opt-Out. All thought that data access decisions should be decided in consultation with patients.

The following key points were agreed:

1. Data use was supported if there were benefits to NHS patients or wider population.
2. Most were not aware of the NHSE National Data Opt-Out.
3. Oversight of data use should be provided by the NHS as the most trusted organisation.
4. Unconsented data use should be limited to what is needed;
5. Patient/public involvement in data access decisions.
6. Transparency of data use. There should be an open dialogue with public and patients.

For direct quotes from the workshop, see Table S2 of the online supplement.

#### The public workshop

This workshop was attended by 30 delegates. 46.7% were based in Birmingham, 43.3% in the wider West Midlands and 10.0% external to the West Midlands. 46.7% identified as male, 50% as female and 3.3% preferred not to say (0% preferred to self-define gender). Median age was 52 years (range 23 - 84) with 20% identifying themselves as from BAME backgrounds. 76.7% had been to an NHS hospital for care but none were undergoing active follow up or treatment.

93.3% were happy for de-identified health data to be used for research and innovation processes, however all agreed there were risks with health data use.

Without knowledge of the patient workshop results, the public workshop agreed the main risks of health data sharing were;

1. Onward data sharing or use without approval
2. Data used against the individual or communities (discrimination and exploitation)
3. Re-identification of the individual
4. Commercial gain from data use (and especially misuse) with no benefit to patients or the UK.

13.3% were concerned about commercial organisations accessing data. All agreed that there should be a searchable record of supported health data access requests. Participants agreed unanimously that patients and the public should be involved in data access processes. Sensitive data or rare conditions were felt more challenging because of the potential consequences or ease of re-identification and that relevant groups should be involved in these data access processes. 76.7% had not heard of the NHS National Data Opt-Out.

The results of the patient workshop were then shared and the group were then asked to consider and agree principles for PIONEER. These were as follows;

1. Health data use with meaningful benefits back to NHS patients and citizens was supported.
2. Data sharing with healthcare providers, academic staff and commercial entities should be considered, as long as there was community support and public awareness campaigns to inform people about how their data was being used.
3. Knowledge of the National Data Opt-Out was low and needed to be improved.
4. Preferably, data should remain in close proximity or within the NHS, with data sharing overseen by the NHS.
5. Data access should be limited to what is needed for a specific project, with agreements about who accesses the data and for how long.
6. There should be patient/public involvement in data access decisions and key advice sought, especially where sensitive fields or rare conditions were included.
7. There should be transparency in how health data is used, including publicly available lists of projects, summaries and benefits.

For direct quotes from the workshop, see Table S2 of the online supplement.

#### Questionnaire results

Demographic details for those that completed the questionnaires are shown in Table 2.

**Table 2:** Demographics of the 308 participants who completed the questionnaires.

Questionnaire results
|  |  | % (N) |  |
| --- | --- | --- | --- |
| **Gender** | Male | 39.3% | (121) |
|  | Female | 58.4% | (180) |
|  | Prefer to self-define | 0% | (0) |
|  | Prefer not to say | 2.3% | (7) |
| **Age** | 13-17 years | 19.2% | (59) |
|  | 18-24 years | 5.5% | (17) |
|  | 25-34 years | 17.2% | (53) |
|  | 35-44 years | 12.0% | (37) |
|  | 45-54 years | 13.0% | (40) |
|  | 55-64 years | 13.3% | (41) |
|  | 65-74 years | 13.0% | (40) |
|  | 75-84 years | 5.2% | (16) |
|  | 85 years or over | 1.6% | (5) |
| **Ethnicity** | White | 70.8% | (218) |
|  | Asian/ Asian British | 14.6% | (45) |
|  | Black/ African/Caribbean/ Black British | 7.8% | (24) |
|  | Mixed/ multiple ethnic groups | 3.2% | (10) |
|  | Other | 2.9% | (9) |
|  | Prefer not to say | 0.6% | (2) |
| **Participant group** | Patient | 22.7% | (70) |
|  | Carer | 8.4% | (26) |
|  | Visitor to hospital | 4.9% | (15) |
|  | NHS Staff | 26.9% | (83) |
|  | NHS Volunteer | 4.5% | (14) |
|  | Public member | 28.9% | (89) |
|  | Unknown | 3.6% | (11) |

Demographics of those completing the questionnaire were broadly comparable to patients who used UHB services but there was less representation of Asian participants than the Birmingham catchment area, based on the 2011 census data. See Figure S2 of the online supplement.

##### Current use of anonymised data

Figure 1 and Table 3 show how respondents thought their healthcare data was currently used. 96.1% thought their data was used for their own healthcare, 71.0% to improve general NHS services and 60.2% used by research by NHS staff. The majority of participants did not think their health data was used by external agencies.

**Figure 1:**
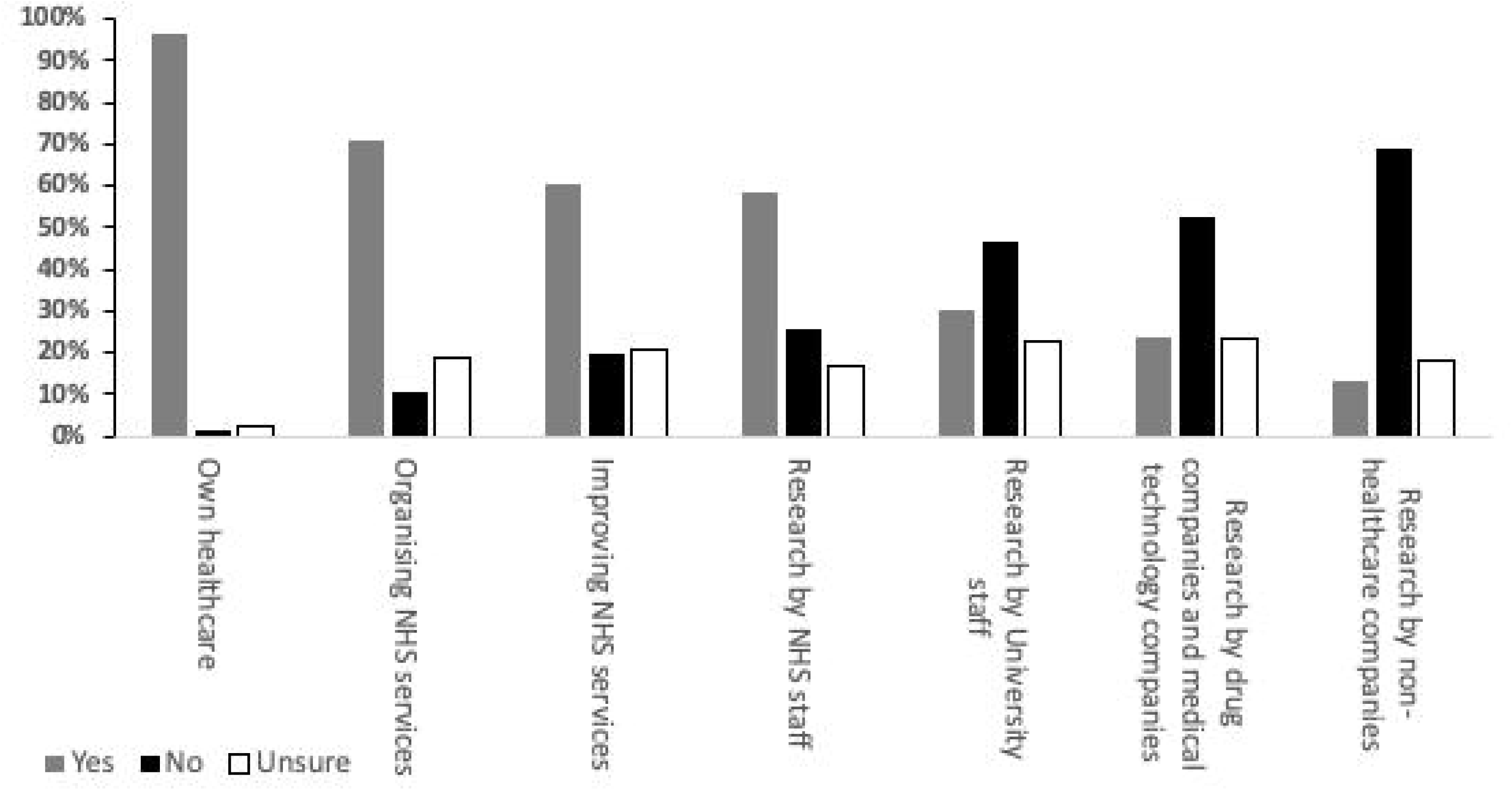
Perception of where healthcare data is currently used. Respondents were asked whether they thought that their health data was currently used for seven purposes, as listed, with a possible answer of yes (shown in grey), no (in black) or unsure (white). All participants answered this question.

**Table 3:** Perception of howanonymised health data is currently used. Respondents were asked whether they believed that their health data was currently used for the seven purposes outlined.

|  | Own healthcare |  |  | Organising services within the hospital |  |  | Projects which improve NHS services |  |  | Research by NHS staff |  |  | Research by university researchers |  |  | Research by drug companies or medical technology companies |  |  | Research by companies who do not provide healthcare products or services |  |  |
| --- | --- | --- | --- | --- | --- | --- | --- | --- | --- | --- | --- | --- | --- | --- | --- | --- | --- | --- | --- | --- | --- |
|  | Yes | No | Unsure | Yes | No | Unsure | Yes | No | Unsure | Yes | No | Unsure | Yes | No | Unsure | Yes | No | Unsure | Yes | No | Unsure |
| **Age (years)** |  |  |  |  |  |  |  |  |  |  |  |  |  |  |  |  |  |  |  |  |  |
| 12-17 | 100% | 0.0% | 0.0% | 54.2% | 18.6% | 27.1% | 47.5% | 33.9% | 18.6% | 57.6% | 32.2% | 10.2% | 33.9% | 47.5% | 18.6% | 25.4% | 47.5% | 27.1% | 8.5% | 66.1% | 25.4% |
| 18-24 | 100% | 0.0% | 0.0% | 64.7% | 5.9% | 29.4% | 52.9% | 29.4% | 17.6% | 70.6% | 17.6% | 11.8% | 47.1% | 41.2% | 11.8% | 29.4% | 58.8% | 11.8% | 23.5% | 64.7% | 11.8% |
| 25-34 | 94.3% | 3.8% | 1.89% | 80.8% | 11.5% | 7.7% | 83.0% | 9.4% | 7.5% | 73.6% | 13.2% | 13.2% | 26.9% | 46.2% | 26.9% | 25.0% | 53.8% | 21.2% | 11.5% | 61.5% | 26.9% |
| 35-44 | 94.6% | 2.7% | 2.7% | 67.6% | 5.4% | 27.0% | 54.1% | 8.1% | 37.8% | 48.6% | 21.6% | 29.7% | 18.9% | 51.4% | 29.7% | 21.6% | 56.8% | 21.6% | 10.8% | 73.0% | 16.2% |
| 45-54 | 87.2% | 2.6% | 10.3% | 73.7% | 10.5% | 15.8% | 68.4% | 10.5% | 21.1% | 60.5% | 13.2% | 26.3% | 34.2% | 31.6% | 34.2% | 26.3% | 31.6% | 42.1% | 23.7% | 52.6% | 23.7% |
| 55-64 | 95.1% | 0.0% | 4.9% | 75.0% | 7.5% | 17.5% | 47.5% | 20.0% | 32.5% | 57.5% | 27.5% | 15.0% | 35.0% | 40.0% | 25.0% | 22.5% | 55.0% | 22.5% | 10.0% | 75.0% | 15.0% |
| 65-74 | 100% | 0.0% | 0.0% | 80.0% | 12.5% | 7.5% | 62.5% | 25.0% | 12.5% | 43.6% | 43.6% | 12.8% | 23.1% | 64.1% | 12.8% | 12.8% | 71.8% | 15.4% | 10.3% | 87.2% | 2.6% |
| 75-84 | 100% | 0.0% | 0.0% | 73.3% | 0.0% | 26.7% | 66.7% | 20.0% | 13.3% | 53.3% | 33.3% | 13.3% | 35.7% | 42.9% | 21.4% | 40.0% | 40.0% | 20.0% | 14.3% | 71.4% | 14.3% |
| 85+ | 100% | 0.0% | 0.0% | 80.0% | 0.0% | 20.0% | 40.0% | 20.0% | 40.0% | 40.0% | 40.0% | 20.0% | 20.0% | 80.0% | 0.0% | 20.0% | 80.0% | 0.0% | 25.0% | 75.0% | 0.0% |
| **Gender** |  |  |  |  |  |  |  |  |  |  |  |  |  |  |  |  |  |  |  |  |  |
| Male | 96.7% | 1.7% | 1.7% | 74.8% | 10.9% | 14.3% | 63.0% | 22.7% | 14.3% | 61.9% | 22.0% | 16.1% | 34.7% | 48.3% | 16.9% | 25.4% | 54.2% | 20.3% | 16.1% | 66.9% | 16.9% |
| Female | 96.1% | 1.1% | 2.8% | 67.9% | 10.7% | 21.4% | 59.2% | 17.9% | 22.9% | 56.4% | 27.9% | 15.6% | 27.7% | 46.3% | 26.0% | 22.5% | 52.2% | 25.3% | 10.7% | 71.2% | 18.1% |
| **Ethnicity** |  |  |  |  |  |  |  |  |  |  |  |  |  |  |  |  |  |  |  |  |  |
| White | 96.3% | 1.4% | 2.3% | 69.6% | 10.3% | 20.1% | 59.1% | 20.0% | 20.9% | 60.3% | 22.0% | 17.8% | 30.5% | 44.1% | 25.4% | 24.4% | 49.3% | 26.3% | 12.7% | 67.9% | 19.3% |
| Asian/ Asian British | 95.6% | 0.0% | 4.4% | 75.6% | 8.9% | 15.6% | 57.8% | 17.8% | 24.4% | 51.1% | 28.9% | 20.0% | 24.4% | 55.6% | 20.0% | 20.0% | 60.0% | 20.0% | 15.6% | 68.9% | 15.6% |
| Black/ African/Caribbean/ Black British | 95.8% | 0.00% | 4.2% | 66.7% | 16.7% | 16.7% | 66.7% | 20.8% | 12.5% | 62.5% | 33.3% | 4.2% | 39.1% | 52.2% | 8.7% | 33.3% | 58.3% | 8.3% | 13.0% | 69.6% | 17.4% |
| Mixed/ multiple ethnic groups | 100.0% | 0.00% | 0.0% | 70.0% | 10.0% | 20.0% | 50.0% | 20.0% | 30.0% | 20.0% | 70.0% | 10.0% | 10.0% | 70.0% | 20.0% | 10.0% | 70.0% | 20.0% | 0.0% | 90.0% | 10.0% |
| Other | 88.9% | 11.1% | 0.0% | 88.9% | 11.1% | 0.0% | 88.9% | 11.1% | 0.0% | 66.7% | 22.2% | 11.1% | 44.4% | 33.3% | 22.2% | 22.2% | 55.6% | 22.2% | 22.2% | 55.6% | 22.2% |
| **Participant group** |  |  |  |  |  |  |  |  |  |  |  |  |  |  |  |  |  |  |  |  |  |
| Patient | 100.0% | 0.0% | 0.0% | 77.9% | 4.4% | 17.6% | 57.4% | 16.2% | 26.5% | 48.5% | 30.9% | 20.6% | 30.9% | 50.0% | 19.1% | 23.5% | 61.8% | 14.7% | 14.7% | 80.9% | 4.4% |
| Patient carer | 96.2% | 0.0% | 3.8% | 76.9% | 15.4% | 7.7% | 46.2% | 30.8% | 23.1% | 42.3% | 46.2% | 11.5% | 30.8% | 57.7% | 11.5% | 19.2% | 57.7% | 23.1% | 16.0% | 68.0% | 16.0% |
| Visiting someone | 93.3% | 0.0% | 6.7% | 66.7% | 13.3% | 20.0% | 60.0% | 6.7% | 33.3% | 60.0% | 13.3% | 26.7% | 26.7% | 33.3% | 40.0% | 26.7% | 40.0% | 33.3% | 0.0% | 80.0% | 20.0% |
| NHS Staff | 94.0% | 2.4% | 3.6% | 78.0% | 11.0% | 11.0% | 77.1% | 12.0% | 10.8% | 73.5% | 18.1% | 8.4% | 30.5% | 48.8% | 20.7% | 20.7% | 57.3% | 22.0% | 11.0% | 69.5% | 19.5% |
| NHS Volunteer | 92.3% | 0.0% | 7.7% | 92.3% | 7.7% | 0.0% | 92.3% | 7.7% | 0.0% | 91.7% | 8.3% | 0.0% | 45.5% | 36.4% | 18.2% | 50.0% | 33.3% | 16.7% | 45.5% | 45.5% | 9.1% |
| Public member | 93.3% | 3.3% | 3.3% | 56.7% | 6.7% | 36.7% | 43.3% | 20.0% | 36.7% | 40.0% | 20.0% | 40.0% | 16.7% | 43.3% | 40.0% | 16.7% | 50.0% | 33.3% | 13.3% | 60.0% | 26.7% |

Comparing the categories of data use, there was no difference in responses by gender or ethnicity. In general, adult volunteers at the hospital thought that health data was currently being used for more purposes than any other group of respondents. Amongst adult respondents, more staff thought data was currently used for research by NHS researchers than those who were patients (73.5% vs 48.5%, p<0.0005).

##### National Data Opt-Out

31.8% were aware of the National Data Opt-Out. Table 4 shows the percentage of respondents aware of the Opt-Out within each demographic group. A higher proportion of women than men were aware of the NHS National Data Opt-Out (36.9% vs 24.2%, p=0.021). A lower proportion of those under 18 years old were aware of the Opt-Out compared to those who were over 18 years old (10.3% vs 36.8%, p<0.0005). NHS staff or volunteers were aware of the Opt-Out compared to those with all other groups (Figure 2, p<0.0005).

**Table 4:** Awareness of NHS National Data Opt-Out scheme. Respondents were asked whether they were aware they could opt-out of their anonymised health data being used.

|  | Yes | No | Unsure |
| --- | --- | --- | --- |
| **Age (years)** |  |  |  |
| 13-17 | 10.3% | 84.5% | 5.2% |
| 18-24 | 29.4% | 64.7% | 5.9% |
| 25-34 | 44.2% | 51.9% | 3.9% |
| 35-44 | 32.4% | 64.9% | 2.7% |
| 45-54 | 46.2% | 48.7% | 5.1% |
| 55-64 | 22.0% | 75.6% | 2.4% |
| 65-74 | 35.0% | 57.5% | 7.5% |
| 75-84 | 56.3% | 37.5% | 6.3% |
| 85+ | 20.0% | 80.0% | 0.0% |
| **Gender** |  |  |  |
| Male | 24.2% | 72.5% | 3.3% |
| Female | 36.9% | 57.5% | 5.6% |
| **Ethnicity** |  |  |  |
| White | 33.6% | 61.3% | 5.1% |
| Asian/ Asian British | 26.7% | 71.1% | 2.2% |
| Black/ African/Caribbean/ Black British | 41.7% | 50.0% | 8.3% |
| Mixed/ multiple ethnic groups | 0.0% | 100% | 0.0% |
| Other | 22.2% | 77.8% | 0.0% |
| **Participant group** |  |  |  |
| Patient | 28.6% | 67.1% | 4.3% |
| Patient carer | 24.0% | 68.0% | 8.0% |
| Visiting someone | 33.3% | 66.7% | 0.0% |
| NHS Staff | 53.0% | 43.4% | 3.6% |
| NHS Volunteer | 71.4% | 21.4% | 7.1% |
| Public member | 16.7% | 83.3% | 0.0% |

**Figure 2:**
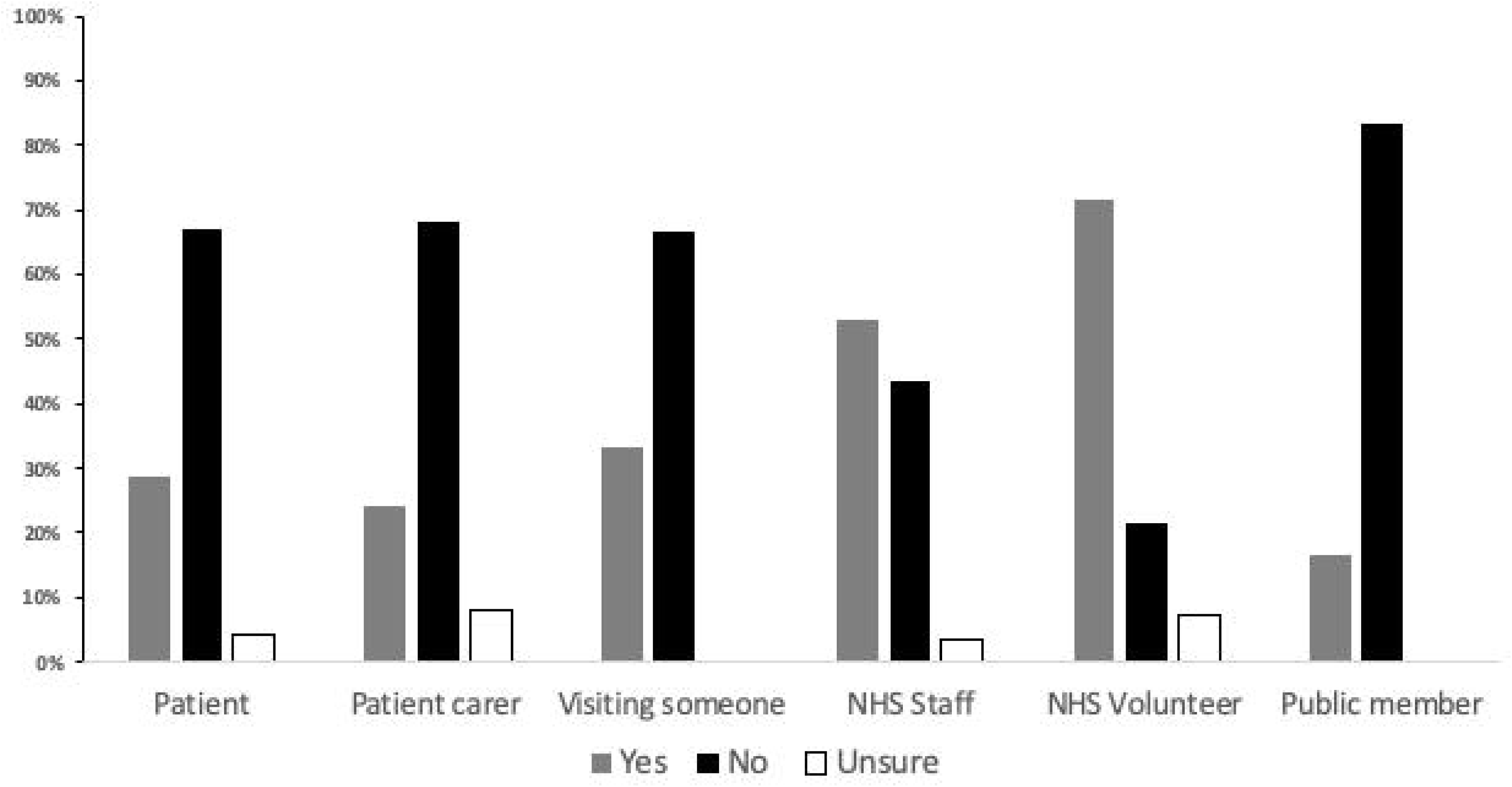
Awareness of NHS data opt-out scheme. Percentage of respondents who were aware of the NHS data opt-out scheme by reason for visiting the hospital or members of the public. All participants answered this question.

##### Acceptable data use

Figure 3 and Table 5 demonstrate the percentage of respondents that would be happy for their anonymised health data to be used for each potential purpose. Three categories were acceptable to more than 90% of respondents: organising NHS services (95.1%); improving NHS services (95.1%); and research by NHS researchers (93.8%). Fewer participants were happy for their data to be used for university researchers than by NHS staff (84.9% vs 93.8%, p<0.0005), although support was high. A higher proportion of those aged 12-17 years old were happy for their anonymised data to be used by drug or medical technology companies compared to those aged 65-74 years or 75-84 years (86.4% vs 55% and 33.3%, p=0.001). Less women than men were happy with data use for organising services (92.3% vs 99.2%, p=0.01), projects that improve NHS services (92.8% vs 98.3%, p=0.031), and research by NHS researchers (91.6% vs 97.5%, p=0.038). There were no other significant differences between responses by gender, ethnicity or reason for visit.

**Table 5:**
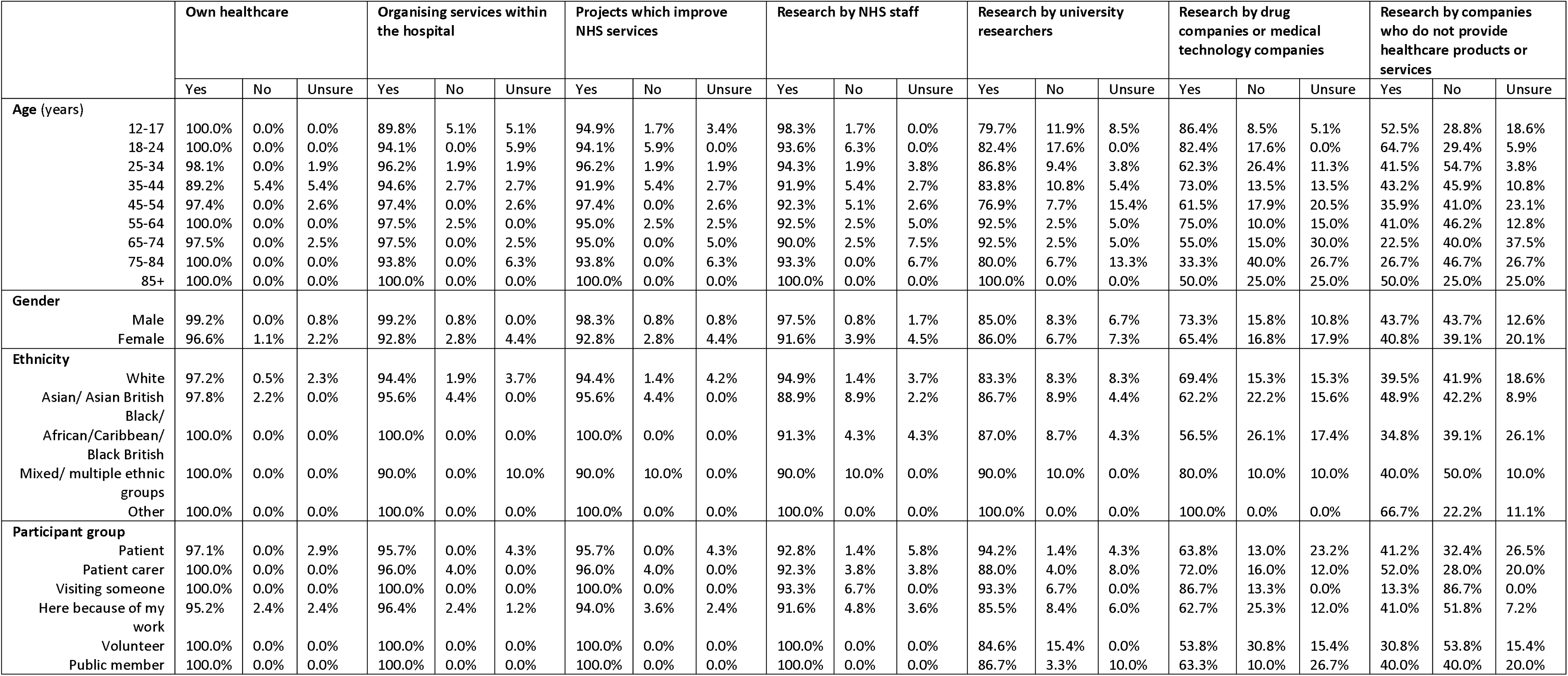
Acceptability of the use of anonymised health data by purpose Respondents were asked whether they would be happy for their anonymised health data to be used for seven purposes, without being asked for their consent.

**Figure 3:**
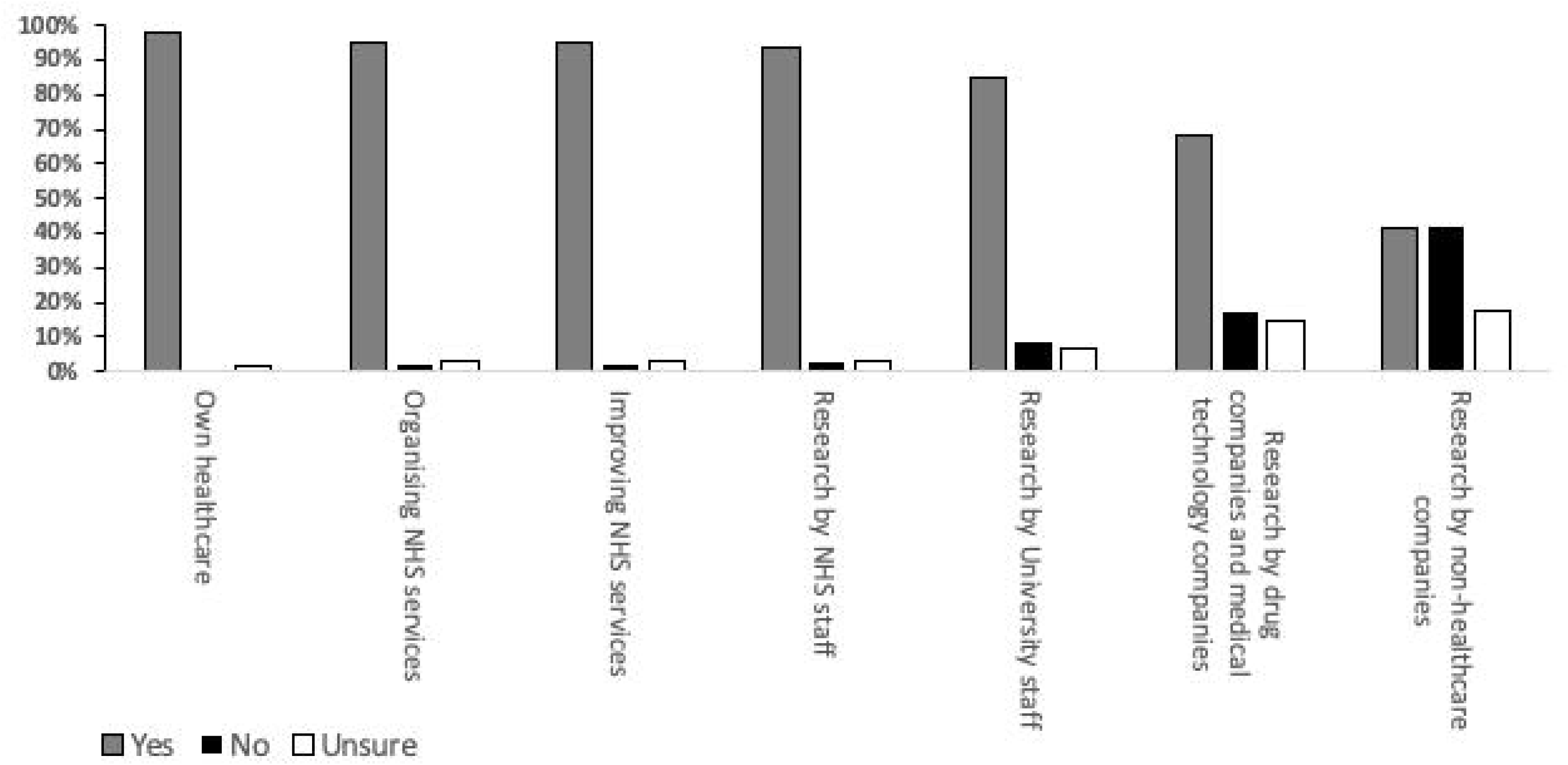
Acceptability of the use of anonymised health data by purpose. Respondents were asked whether they would be happy for their anonymised health data to be used for seven purposes. All participants answered this question.

##### Current use compared to acceptable use

Responses for each category were assessed, comparing how respondents thought data was currently used and whether they would be happy for the data to be used. For all categories, a significantly higher proportion of respondents said they would be happy for their data to be used than thought it was currently used for this purpose (Table 6).

**Table 6:** Comparison of proportion of respondents who think their health data is currently used for each suggested purpose, compared to the proportion that would be happy for their data to be used in this way.

|  | Think is currently used for this reason | Would be happy for use for this reason | p value |
| --- | --- | --- | --- |
| Own healthcare | 96.1% | 97.7% | 0.244 |
| Organising services within the hospital | 71.0% | 95.1% | <0.005 |
| Projects which improve NHS services | 60.2% | 95.1% | <0.005 |
| Research by NHS staff | 58.1% | 93.8% | <0.005 |
| Research by university researchers | 30.2% | 84.9% | <0.005 |
| Research by drug companies or medical technology companies | 23.8% | 68.4% | <0.005 |
| Research by companies who do not provide healthcare products or services | 13.0% | 41.3% | <0.005 |

#### Feedback groups

The results of the questionnaires were presented and discussed with the patient and public workshop attendees. There was agreement that health data use was broadly supported. As knowledge of the NHS National Data Opt-Out was low, people did not have the opportunity to exercise their right to opt out through a potential lack of awareness. The seven principles to guide unconsented health data use described above were ratified without change.

### Discussion

Different countries operate different legal frameworks for health data access, with the UK choosing a National Data Opt-Out. Recent questionnaires suggest variable support for data access, but this may depend on the population asked. Few people have asked young adults and the views of frequent healthcare users may differ from those without chronic illnesses. There may be regional differences related to where the data is stored and which organisation is Data Controller. The PIONEER Hub contains health data from the West Midlands and so the opinions of local patients and citizens were sought. We describe the initial phases of a program of work to explore these themes.

In general, there was support for the use of anonymised health data for secondary purposes by NHS, academic and commercial organisations, providing there was patient and public involvement and engagement in data-sharing decisions and outcomes. Only research by non-healthcare commercial organisations receiving less than 50% support. The reported support for health data use in the current paper are much higher than reported recently^(12)^. The reasons for the disparity are unclear. This might reflect differences in the population questioned or the inclusion of patients and visitors to hospital and NHS staff.

Awareness of the NHS National Data Opt-Out system was low including in NHS staff. This suggests that increased education is needed for the public, patients and carers, as well as those working in healthcare.

Previous surveys also suggest that public citizens have greater confidence in data use for research conducted through the NHS than by pharmaceutical companies^(21)^. In our study, the NHS was consistently identified as the most trusted partner to hold data or make decisions on health data use, mirroring the findings of a recent OneLondon event^(22)^ and previous research^(11,23)^

The results from our study are in keeping with those recently published from Understanding Patient Data, which described a majority of people believing the public should be involved in decisions about how NHS data is used and that benefits from health data partnerships should be shared across the NHS^(24)^

The principles of data minimisation (access only to what data was needed and no more, by only those who needed to access the health data) viewed favourably by our participants has also been highlighted in previous engagement events^(22)^ and may help to increase acceptability of data use for secondary purposes^(25)^.

Those aged 13 years or older were included here as individuals can choose to use the NHS data National Data Opt-Out from the age of 13. Our results suggest those under 18 may be more accepting of anonymised data use by pharmaceutical or medical technology companies than older adults, and that they had lower awareness of the NHS data National Data Opt-Out that adults (10% aware). A previous study of the perception of electronic health records found that 60% of young people did not understand how healthcare records could be beneficial to research, which improved after education^(26)^.

This study has limitations. The sample size was limited and should not be extrapolated across other populations. Further work is needed to understand if perceptions of health data use differ across different groups or communities. Workshops allowed for a more in-depth assessment of patient and public perceptions, but did not fully explore why participants would not be happy for their data to be used for specific purposes.

### Conclusion

There remains both support and concerns with secondary use of health data. However, concerns can be reduced by increasing public awareness of data use and public choice, the involvement of patient and public voices in health data access decisions, public good at the heart of data sharing, proximity of data access decisions to the NHS and the well-established principles of data minimisation.

## Data Availability

Structured data is available from the PIONEER Hub upon reasonable request.  Please contact the corresponding author for details.

## Author contribution

CA, BC, KD, ES, GP, EM, MO’H, CM, AI, CC, ML, SM, SG, SP, JA, GG, RD, AR, IA, HF designed the study, facilitated the public and patient workshops and contributed to the manuscript. CA, BC, SG, ES analysed data. CA and ES wrote the first version of the manuscript. CA, BC, KD, ES, SG, HF, GP, MO’H, CM, AI oversaw data collection. All authors approved the final version.

## Conflicts of Interest

CA, BC, KD, GP, EM, MO’H, CM, AI, CC, ML, SM, SG, SP, JA, RD, AR, HF have no conflicts of interest. GG reports funding from HDR-UK. AKD reports funding from HDRUK, The Wellcome Trust and Fight for Sight. ES reports funding from the Wellcome Trust, MRC, HDR-UK, Alpha 1 Foundation (A1F), British Lung Foundation and NIHR.

## Acknowledgements

This work was supported by PIONEER, the Health Data Research Hub in acute care which is funded by Health Data Research UK (HDR-UK). HDR-UK is an initiative funded by UK Research and Innovation, Department of Health and Social Care (England) and the devolved administrations, and leading medical research charities. We would like to acknowledge the contribution of all staff, key workers, patients and the community who have supported our hospitals and the wider NHS at this time.

## Notes

### Competing Interest Statement

CA BC KD GP EM MOH CM AI CC ML SM SG SP JA RD AR HF have no conflicts of interest.  GG reports funding from HDR-UK. AKD reports funding from HDRUK, The Wellcome Trust and Fight for Sight. ES reports funding from the Wellcome Trust, MRC, HDR-UK, Alpha 1 Foundation, British Lung Foundation and NIHR.

### Clinical Trial

Not applicable

### Author Declarations

PIONEER is an ethically-approved research database and analytical environment (East Midlands Derby Research Ethics 20/EM/0158). This PPIE work was conducted following ethical approval from the University of Birmingham Ethical Committee (reference ERN_20-0118).

